# Rapid increase of SARS-CoV-2 seroprevalence during the 2020 pandemic year in the population of the city of Tirana, Albania

**DOI:** 10.1101/2021.02.18.21251776

**Authors:** Genc Sulcebe, Alban Ylli, Fabian Cenko, Margarita Kurti-Prifti

**Author notes:** Corresponding Author: Alban Ylli.

## Abstract

**Introduction:** While the identification of anti SARS-CoV-2 antibodies has been used to measure the hidden circulation of the COVID-19 in communities, there are few publications on the dynamics of SARS-CoV-2 seroprevalence during both waves of 2020. This study provides original data about the change in proportion of individuals showing immune response to COVID-19 between beginning of July and end of December 2020.

**Methods:** The study was conducted in two rounds, 27 June −3 July, and 21-28 December 2020, using two independently selected samples of individuals 20-70 years old. Study participants were randomly selected from lists of the inhabitants of the catchment communities of four primary health care centers in Tirana City. Serological testing was performed by an ELISA method which determines IgG class antibodies anti S1 protein of SARS-CoV-2 virus. The validity of the method was tested in a sample of blood donor’s sera of 2018.

**Results:** The proportion of individuals classified as seropositive during the first round, in early July was 7.5% (95% CI: 4.3% −10.7%). The proportion rose sharply in the second round, by late December 2020, reaching 48.2% (95% CI: 44.8% −51.7%). The same increasing pattern was observed in all studied categories. No statistical significance was found between men and women and between age categories. The prevalence of seropositive individuals was always significantly higher among those who reported symptoms and those who had done the molecular test.

**Conclusion:** The ratio of total infected cases over confirmed cases was estimated to be higher than 10 to 1 in Albania. The rapid increase in SARS-CoV-2 seroprevalence observed in Tirana City may have been facilitated by a number of factors, including the very low infection exposure during the period March -May 2020, and the consecutive high susceptibility in population. Despite the observed high seroprevalence, one month after the study, COVID-19 incidence continued to increase in Tirana.

## Introduction

Monitoring the prevalence of COVID-19 infection is an important tool for understanding the current epidemiological situation of this infection in the population and consequently for taking the appropriate measures in order to prevent and limit the impact of the epidemic (1, 2). Testing and tracing the SARS-CoV-2 infection through molecular or antigenic testing, which detect the presence of the virus, undoubtedly play an important role for recognizing the incidence of the infection (3, 4). However, these examinations have their limitations in terms of the accurate assessment of the true infection rate in the population. This is due to the fact that pre-symptomatic and asymptomatic individuals may go undetected through direct viral testing (5). In fact, many individuals with mild or moderate forms of the disease, for various reasons either could miss viral examinations or could obtain negative results due to delayed testing (4, 6).

Serological testing, carried out through the determination of the presence of anti SARS-CoV-2 antibodies in the serum, provide accurate and relatively reliable information on specific immunization against the virus (7, 8, 9). Thereby, serosurvey studies help to show the true prevalence of COVID-19 disease in the population, as they provide retrospective information on the cumulative infection rate in the population (10, 11).

SARS-CoV-2 seroprevalence studies are mainly based on identifying the presence of IgG antibodies anti Spike (S) or Nucleocapsid (N) viral proteins of SARS-CoV-2 virus in human sera (12). The data provided by these studies have helped to assess the real prevalence of COVID-19 infection in the general population during the spring and summer months of 2020 in many countries (13, 14). However, at least in Europe, there are still very few publications on the dynamics of SARS-CoV-2 seroprevalence during the second wave of autumn and winter 2020.

Tirana, the capital of Albania, retains about a third of the country’s urban population (15). Therefore the information on the prevalence of COVID-19 in this city is also useful in terms of estimating the prevalence of this infection throughout the country. No studies on COVID-19 seroprevalence in the general population have been conducted or published until December 2020 in Albania, which explains the importance of the results produced by a two-round seroepidemiological survey carried out over two periods during 2020 (the end of June and the end of December 2020).

The main hypothesis of the study was that the intensive circulation of SARS-CoV-2 virus in the city of Tirana, after the end of the initial lockdown in early June 2020, would lead to a rapid increase of the seroprevalence of anti-SARS-CoV-2 antibodies in the following months. Furthermore, this prevalence was expected to be higher than the proportion of individuals formally diagnosed with COVID-19 through direct viral testing. The seroprevalence is analyzed in relation to several characteristics to identify differences in various population categories. Finally, the information obtained from this study is also triangulated with data from other sources to estimate the COVID-19 prevalence in the entire Albanian population for the same time period covered by the survey.

## Material and Methods

### Sampling methodology

A two-level systematic sampling methodology was applied to ensure the generalization of results into the larger adult population of the city of Tirana.

In the first level, out of the 11 public health centers (HCs) that cover the city of Tirana, four HCs were randomly selected; respectively HC number 3, HC 7, HC 8 and HC 9, whose catchment population is about 258 000 inhabitants. The selected HCs cover strictly urban population of Tirana. In the second level, the physicians and head nurses of the four HCs were instructed to randomly select a number of individuals aged 20 to 70 years old from the family physician registries. Each family physician, as part of the selected HCs, has an electronic list of the inhabitants of their catchment communities. The list of the resident population is updated yearly according to the contract each HC signs with the Compulsory Health Insurance Fund (the only public health services buyer in the country).

The study was conducted in two rounds by means of two independently selected samples; 300 individuals invited during the time interval 27 June to 3 July 2020 and 900 individuals invited during the time interval 21 to 28 December 2020. The smaller sample in the first round was due to the limited number of available serological tests at that moment.

Each selected HC was instructed to randomly select 75 individuals aged 20 to 70 years old in the first round and 225 individuals in the second round. Materials for blood sampling, the sample forms to be completed for each individual tested, and relevant instructions were distributed to each family physician of the selected centers in the weeks previous to each study round. Medical and nursing staff were trained to conduct the study. The selected individuals were invited to participate in the study by phone calls and their prior approval for the relevant blood sampling and laboratory testing was ensured.

Blood samples taken from each health center were transported within 3 hours to the Laboratory of Immunology at the University Hospital Center of Tirana to perform laboratory procedures for serological testing. Additional data were collected from each participant such as demographic (age, gender, residence), anamnestic data on previous COVID-19-like symptoms, and/or SARS-CoV-2 molecular testing carried out during the months preceding their participation in the study. The current blood samples for each round were smaller than the number of the invited individuals due to a small number of refusals. However, the refusal rate did not exceed 10% (266 vs. 300 for the first round, and 817 vs. 900 for the second round).

### Serological testing of IgG class anti-S1-CoV-2 antibodies

Serological testing of all blood samples was performed by an ELISA method using a commercially available diagnostic kit which determines IgG class antibodies anti S1 protein of SARS-CoV-2 virus (IgG anti S1-SARS-CoV-2 ELISA, Euroimmun, Luebeck, Germany). All laboratory procedures were carried out according to the manufacturer’s instructions.

To validate the specificity of the IgG anti S1-SARS-CoV-2 ELISA kit, this kit was used to test the sera of 100 blood donors obtained from the Central Blood Bank of Tirana, which were randomly selected from the year 2018, when SARS-COV-2 is assumed not to be present. The diagnostic sensitivity of this diagnostic kit was checked with a similar kit from the Euroimmun company, which uses the nucleocapsid protein of the virus (IgG anti NCP-SARS-CoV-2) as antigen. For this purpose, 92 sera tested for IgG anti S1-SARS-CoV-2 were also tested with the IgG anti NCP-SARS-CoV-2 ELISA kit.

For both diagnostic kits used, the results were evaluated semi-quantitatively by calculating the ratio of the optical density extinction of the sample over the extinction of the calibrator as following the manufacturer’s recommendations. The values above 1.1 are considered as positive, those between 0.8 to 1,1 as borderline and those inferior to 0.8 as negative. In our study, only the serum samples with values above 1.1 were considered as seropositive.

All raw data have been statistically analyzed using the SPSS 20 package programs.

## Results

In the first round of survey, between June 27 and July 3, 2020, 266 participants were enrolled and in the second round, during the period 21 to 28 December 2020, 817 participants were enrolled. There were very few missing data in the final database. Table 1 is organized according to the two study rounds. For each round the following data have been provided: demographics, COVID-19 self-reported symptoms, previous molecular testing, and serology (including CI: Confidence Interval).

**Table 1.**
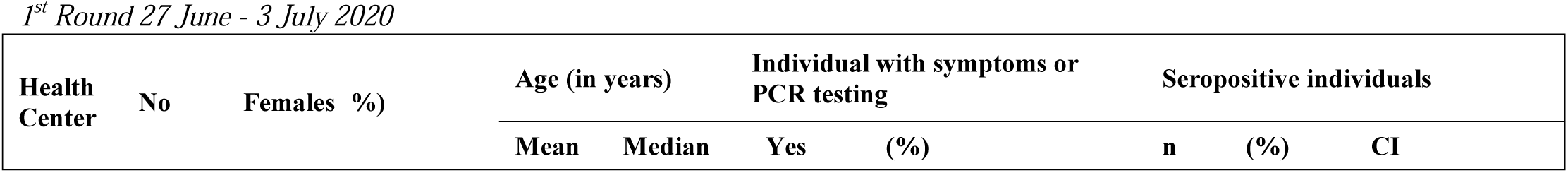
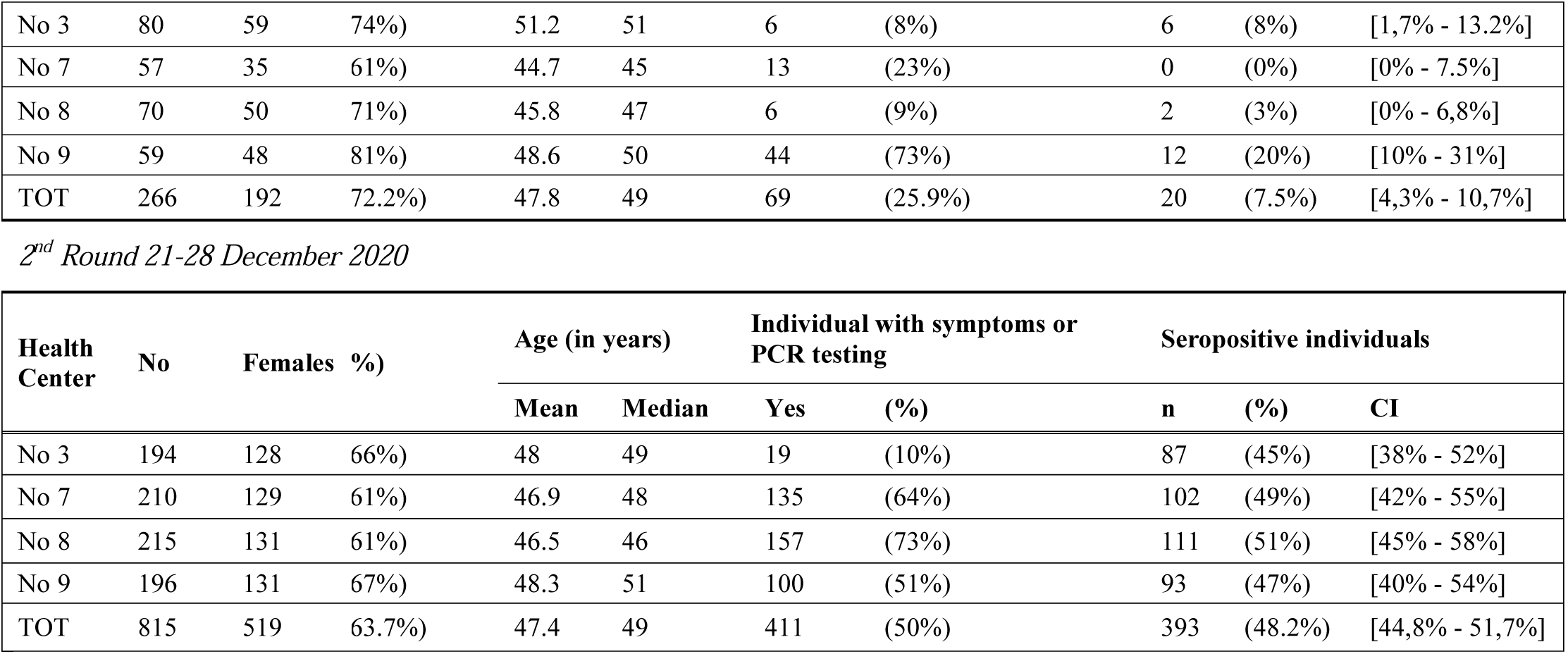
General demographic, clinical and serology data.

Only around 28% of participants were male in the first study round, but the proportion of male participants rose to over 36% during the second round, approaching that of the normal population (49%). The 10 year age-groups distribution in the sample was found similar to the general Albanian population aged 20 to 70 years old (15).

The proportion of individuals classified by the test as IgG anti S1-SARS-CoV-2 seropositive (from herein called simply *seropositive*) during the first round of the study was 7.5%. The proportion rose sharply in the second round, reaching 48.2% (or around 6.5 higher compared to the first-round result). The same increasing pattern was observed in all studied categories (HCs, age and sex).

There were 58 individuals enrolled by chance in the samples of both study rounds. In the first study round only 4 or 6.9% of these individuals resulted seropositive, while in the second round 31 or 53% of these individuals resulted seropositive. Interestingly, the point prevalence of the seropositivity in the subsample of these “followed up participants” was very similar to the rate found in both total study samples of late June and late December (Chi Square = 0.1; p-value = 0.6).

Table 1 shows that the significant difference in the seroprevalence rate between selected HC catchment areas observed during the first study round disappears in the second round. In late December, the proportion of seropositive individuals ranged from 45% to 51% between four HC-based subsamples, but there was no significant difference amongst them (Chi Square rate =1.8, p = 0.6).

The prevalence of the seropositive individuals for each 10-year subgroup at the second round varies from 44% till 52%, but no statistical significance was found (Chi Square test = 2.824, df = 4, p = 0.56). Likewise, no statistical significance was found in sex-based seroprevalence analysis; 51% and 44% for women and men respectively (OR = 0.76, CI [0.6 −1.01]).

Table 2 shows that during the first round, 25.1% (67/266) of participants reported to have had symptoms similar to those to COVID-19 during previous 3 months (95% CI: 20.3% −30.7%). and 8.2% of participants (22/266) reported to have done the molecular test for SARS-CoV-2 (95%CI: 5.5% −12.2%). Conversely, in the second round more than 50% (407/807) of participants reported symptoms during previous 8 months (95% CI: 46.4% −53.2%), and 12% (97/807) reported to have done the SARS-CoV-2 specific molecular test (95% CI: 9.8% −14.2%).

**Table 2.**
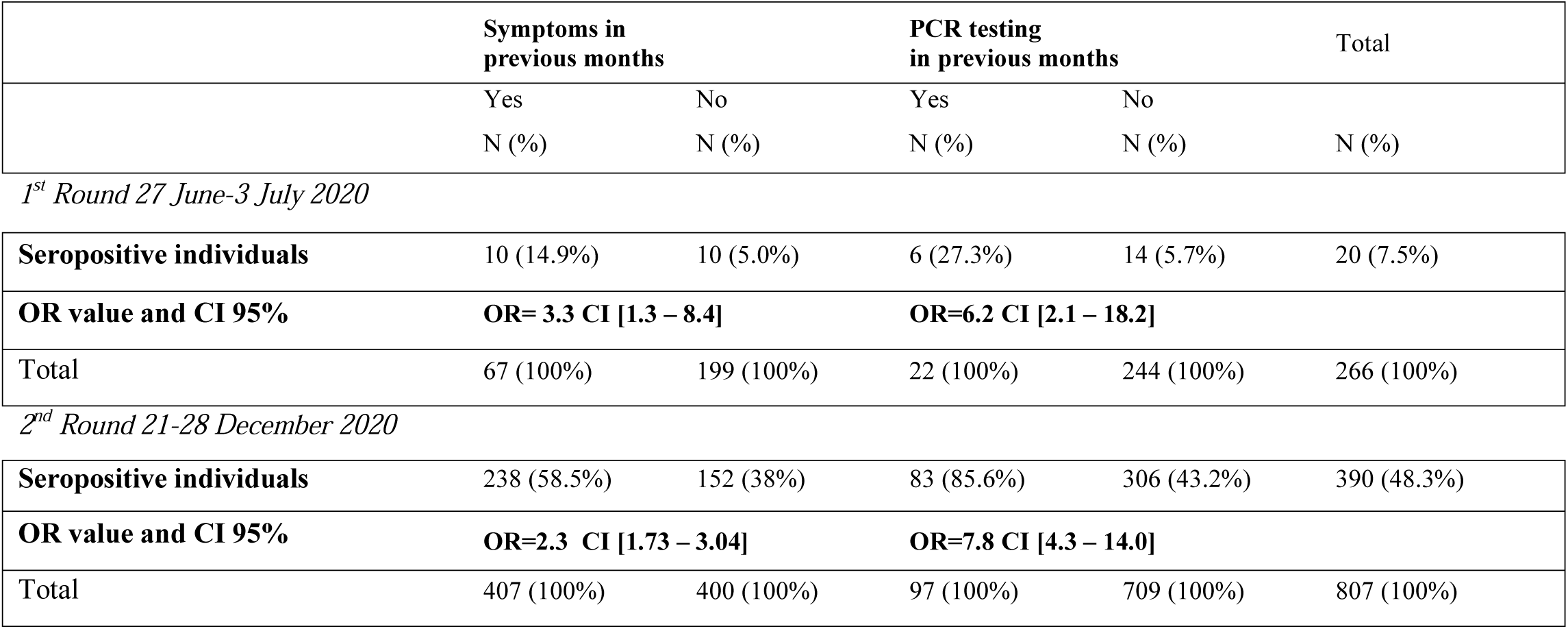
Distribution of seropositive results according to reporting of previous symptoms and previous molecular test *(into two study rounds: 27 Jun-3 July and 21-28 December)*

In both rounds of study, the prevalence of seropositive individuals is always significantly higher among those who reported symptoms and those who had done the molecular test. In first and second study round respectively, the odds to be positive in serological test for those who report previous symptoms were 3.3 and 2.3 higher compared to those who did not report symptoms. The odds to be serologically positive is even higher for those reporting to have had a molecular test; 6.2 and 7.8 higher compared to those who didn’t have any previous test, in the first and second study round respectively.

Among the 100 blood donor sera from the blood bank collection of the year 2018, two sera tested positive (2%) for anti S1-SARS-CoV-2 IgG antibodies. Both sera were checked for anti nucleoprotein (N) SARS-CoV-2 IgG antibodies with diagnostic kits from two other manufacturers and both resulted positive also with these tests.

In order to compare the diagnostic sensitivity of the IgG anti S1-SARS-CoV-2 diagnostic test that we used throughout this study, 92 selected sera that were tested for these antibodies during the second round of study were also tested for IgG anti NCP-SARS-CoV-2 antibodies. All 42 IgG anti S1-SARS-CoV-2 positive sera also resulted positive for IgG anti NCP-SARS-CoV-2. Meanwhile, among the 50 IgG anti S1-SARS-CoV-2 negative sera, 6 tested positive for IgG anti NCP-SARS-CoV-2. From this comparative testing we can conclude that in order to determine the seroprevalence of SARS-CoV-2 infection, anti NCP-SARS-CoV-2 IgG antibodies have a 12.5% higher sensitivity than IgG anti S1-SARS-CoV-2 in our study.

## Discussion

In this report we present data on IgG anti-S1-SARS-CoV-2 seroprevalence in a sample of individuals representing the population aged 20 to 70 years resident in Tirana city, Albania. This study, conducted in two rounds, at end of June 2020 and the end of December 2020, reflects the dynamics of the seroprevalence in this population during the 2020 COVID-19 pandemic year. The two rounds of the study demonstrate a much higher rate of SARS-COV-2 circulation in the community than the number of confirmed COVID-19 cases. Furthermore, as hypothesized, the SARS-COV-2 seroprevalence rose on average 6.5 times in a six-month interval from the end of June to the end of December 2020. This period corresponds to a continuous increase of daily COVID-19 cases reported by the health system, peaking in November-December 2020 (16).

Albania, like many countries in East Europe did not experience the intensity of the COVID-19 crisis, which overwhelmed many Western countries during the first wave of pandemic in Winter-Spring 2020. The drastic public health measures were timely and effective. (17). While the number of daily cases was kept quite low initially in Albania, with the reopening phase during spring-summer 2020, the cumulative incidence and case fatality rate began to rapidly increase.

On 30 June 2020, a total of 2535 COVID-19 cases were reported in the country, and only 62 deaths. Around half of the cases were reported from Tirana district. By the end of December, the time of second round of this study, the COVID-19 diagnosed cases and deaths were respectively 55380 and 1134, again with more than half of the cases reported from Tirana district (18). Overall, the increasing trend of seroprevalence reported in this study mirrors the trend of reported COVID 19 cases and deaths during the period June-December 2020 in Albania.

Overrepresentation of women in the study samples may reflect the underutilisation of public health care by men in Albania, which has been previously reported (19). A similar finding has been reported by a seroprevalence study in Greece (20). The slightly higher SARS-COV-2 seroprevalence among women, although not statistically significant, coupled with their overrepresentation in the study sample, may have influenced a slightly higher result than the real seroprevalence in the general population in the first study round. Additionally, in the first study round, an overrepresentation of individuals in population who had done a molecular testing is noted, and this fact may have also contributed to a result higher than the real population seroprevalence. Nonetheless, the proportion of participants who had undergone the molecular COVID-19 test in the second round of the survey sample is very similar to the indicator “tests per population”. By 15 December the test per population rate in Tirana was around 20%, which is higher than the rate found in the study sample (12%). This shows that there was no selection bias towards those individuals at higher risk for the disease. Additionally, the age distribution of participants in the sample reflects the proportion of Albanian population age categories, with only individuals aged 20-30 years old being slightly under-represented (21). The study did not find important differences in SARS-COV-2 seroprevalence among age-groups, a finding that has been reported by other serological studies (22).

It is worth mentioning that a 2 % IgG anti-S1-SARS-CoV-2 positivity in the blood donors’ sera from the year 2018 was found. These samples were positive for IgG antibodies anti both S (spike) and nucleoprotein (N) viral proteins. Most likely this positivity reflects a cross-reactivity with human coronaviruses that are often encountered in seasonal infections in all populations (23). Other studies have also found similar positivity in the sera of group-individuals tested before 2019 (24, 25). The seroprevalence results should therefore be interpreted within the limits of the test specificity, keeping in mind a potential of around 2% IgG false positivity in both rounds of survey. On the other hand, it should be underlined that anti SARS-CoV-2 IgG is not the only indicator of immune response against the virus. Hence, there may be suspected more individuals in the population, potentially exposed to COVID-19, than this study can document.

39 % of all seropositive individuals in this study reported no COVID-19 like symptoms. This proportion corresponds to other studies that have reported this value in the range from 32,4 % in England to 57,2 % in Iran (2, 26, 27).

Based on the study results and the analyses presented above in this section, it can be concluded that, when compared to the proportion of cases confirmed by the health system, the proportion of SARS-COV-2 infected cases in the population is many times higher. The lack of detailed data related to distribution of COVID-19 reported cases by municipalities in Albania and the lack of recent data on the resident population of urban Tirana, makes difficult the precise estimation of the number of individuals who have developed specific IgG anti COVID-19, both in Tirana and in the whole country. The same is true for the estimation of the precise ratio of infected cases over confirmed cases. Nevertheless, secondary data based on cumulative reported cases, active cases and deaths, retrieved from daily official reports (18) help to roughly estimate that almost half of the total number of COVID-19 cases diagnosed in Albania have been reported from Tirana district. Carefully extrapolating study results into the general population of Tirana City and Tirana District, and comparing them to the number of confirmed COVID-19 cases in Tirana, allows for an approximate estimation of the ratio of total infected cases over confirmed cases. The real number of individuals in Tirana City showing immune response to SARS-COV-2, should have been between 250000 −300000, at a time when confirmed cases in the same population were less than 25000. The ratio of total infected cases over confirmed cases was then expected to be higher than 10 to 1. A similar ratio was already reported by other surveys using similar methodology (7, 20, 28). Applying the estimated ratio at a national level in late December, it could be generated a figure of at least 600,000 infected cases at that time in Albania, This figure consist of 20% to 25% of the total resident population of Albania (15).

The ratio estimated here should be generalized with caution, as it was generated in specific circumstances, such as an intense community circulation of SARS-Cov-2 in a dense urban population. The ratio may fluctuate within a broad range in other conditions. Numerous studies on the seroprevalence of anti-SARS-CoV-2 antibodies conducted worldwide during the COVID-19 pandemic give different results depending on the urban density, the measures taken to limit the transmission of the virus in the community or the stage of the pandemic. In Europe, the seropositivity has been reported from 5% in Spain in April −May, to 6.7% in France in June 2020 (13, 29). In the USA the prevalence of seropositivity was 19.1% in New York City in the month of April (30), and at the same time it was 4.65% in California (31). High seropositivity rates (respectively 22%, 42% and 66%) have been reported in April 2020 in Guilan, Iran and in July 2020 both in Bergamo, Italy and in Manaus, Brazil (32, 33, 34).

However, to our knowledge, there are no peer reviewed publications documenting changes in seroprevalence between the first and second wave of the COVID 19 pandemic in Europe. A recent preliminary report from a UK survey showed an increase of only two times between June and December (35).

The very strong increase in seroprevalence observed in Tirana city may have been facilitated by a number of factors. Easing of social distancing measures, along with the very low infection exposure during the period March-May 2020, and the consecutive high susceptibility in population, may be some of them. Further studies are needed to link the trends of seroprevalence of anti-SARS-CoV-2 antibodies during the second half of 2020 with COVID-19 mortality rates and excess mortality during the same period.

The data reported in our study are of interest for evaluating the measures for controlling COVID-19 infection during 2020, and planning the needs for the vaccination campaign in 2021. It is important to note that the high seroprevalence in December in Tirana was not followed by any decrease in infection rates, in January 2021. A similar phenomenon is observed in other populations with very high infection rate such as Manaus, Brazil (36). A number of factors, including newly discovered variants of SARS-COV-2, can influence the infection rate during 2021. In conclusion, the recommended public health measures must continue to be observed and the vaccination program must be strengthened to prevent as many deaths as possible.

We should also note that our study has some limitations. The sample size of the population especially in the first phase was rather limited and the percentage of seropositivity was calculated with a somewhat wide confidence interval (4.3% to 10.7%). Our study was carried out by the determination of anti-Protein S1 SARS-CoV-2 IgG antibodies. As we observed in our comparative sensitivity study, anti-nucleocapsid (N) SARS-CoV-2 IgG antibodies have at least 10% higher sensitivity than anti S1-SARS-CoV-2 IgG for COVID-19 testing, a finding also reported from other sources (8, 12, 37). From this finding we can infer that if we had used the anti-nucleocapsid (N) SARS-CoV-2 IgG testing, the seroprevalence rate in our population sample could be even higher than that which was obtained by IgG anti S1-SARS-CoV-2 IgG testing. The seroprevalence results, especially in the case of proportions fewer than 5% and especially in the first round of the study, should be interpreted with caution and always within the limits of test validity. Also, in our study we have investigated the seroprevalence of SARS-CoV-2 infection only in Tirana city residents aged 20 to 70 years who have access to public health services.. The fact that the sample does not include those who do not use primary health care or do not collaborate with their health workers should also be taken into consideration when applying the results in general population.

## Data Availability

All data are fully available without restriction. Yet, data cannot be shared publicly because of they contain the names of participants.
Data can will available to editors and reviewers

## Acknowledgements

Authors acknowledge the essential contribution of Dritan Ulqinaku, Bajram Dedja, Bruna Shiroka, Blerta Berberi, Bujar Mema, Ilirjan Gjyzari, and Irena Seferi.

## Ethical considerations

The methodology and ethics of both rounds of the study were approved by the committee of the Academy of Sciences of Albania

